# Exploring the neuronal and systemic physiological contributions to spontaneous cerebral fluctuations – Insights from functional MRI in The Maastricht Study

**DOI:** 10.1101/2023.12.06.23299117

**Authors:** L.W.M. Vergoossen, J.F.A. Jansen, M.T. Schram, J.J.A. de Jong, M.P.J. van Boxtel, J.P.H. Reulen, A.J.H.M. Houben, M.M.J. Greevenbroek, D. Huybrechs, W.H. Backes

**Affiliations:** Department of Radiology & Nuclear Medicine, Maastricht University Medical Center+ (MUMC+), Maastricht, the Netherlands; MHeNs School for Mental Health and Neuroscience, Maastricht University, Maastricht, the Netherlands; School for Cardiovascular Disease (CARIM), Maastricht University, Maastricht, the Netherlands; Department of Internal Medicine, Maastricht University, Maastricht, the Netherlands; School of Nutrition and Translational Research in Metabolism (NUTRIM), Maastricht University, Maastricht, the Netherlands; Department of Psychiatry and Neuropsychology, Maastricht University, Maastricht, The Netherlands; Department of Clinical Neurophysiology, Maastricht University Medical Center+ (MUMC+), Maastricht, the Netherlands; Department of Electrical Engineering, Eindhoven University of Technology, the Netherlands; Department of Computer Science, K.U. Leuven, Leuven, Belgium

**Keywords:** Resting-state functional MRI, BOLD signal, Wavelet transformation, Cardiometabolic risk factors

## Abstract

**Objective:** Functional MRI (fMRI) is sensitive to changes in the blood oxygen level-dependent (BOLD) signal, which originates from neurovascular coupling, the mechanism that links neuronal activity to changes in cerebral blood flow. Isolating the native spontaneous neuronal fluctuations from the BOLD signal of the resting-state is challenging, as the signal induced by neuronal activity represents only a small part of this signal. Furthermore, many other non-neuronal (systemic) oscillations contribute, such as from the cardiovascular and respiratory system with frequencies partly overlapping those of the spontaneous fluctuations. The objective of this study is to investigate to what extent various systemic physiological signals are associated with the measured BOLD signal, in particular the frequency interval pertaining to the spontaneous cerebral fluctuations (10-100 mHz). Additionally, we investigate whether these associations were independent of cardiometabolic risk factors.

**Methods:** Within the population-based Maastricht Study, 3T resting-state functional MRI and physiological measures, covering cardiac, respiratory, myogenic, neurogenic, and endothelial activity, were acquired (n=1,651, 48% woman, aged 59±8 years). As both neuronal and non-neuronal physiological signals contain frequencies that vary over time, a wavelet transformation (WT) was used. Time-series were decomposed into seven wavelet subbands, and for each subband, the energy of the BOLD signal was calculated. Multivariable linear regression analysis was used to investigate the association of the physiological measures, in particular cognitive function, with the wavelet energy per subband, independent of cardiometabolic risk factors.

**Results:** We found that physiological measures were associated with the energy of certain frequency subbands of the spectrum of the measured fMRI signal. Both cognitive performance and blood pressure variations, as measures of neurogenic and myogenic activity respectively, were associated with the energy of the frequency subband 3 (31.2-62.5 mHz). Furthermore, cardiac and respiratory activity were associated with the energy of the high frequency subband 1 (>125 mHz), and endothelial activity with the energy of low frequency subbands 6 and 7 (<10 mHz). Part of these associations were dependent on cardiometabolic risk factors.

**Conclusion:** We found an association between myogenic and neurogenic activity and the frequency specific BOLD signal. Our findings highlight the strong intertwining of neuronal, vascular, and cardiometabolic activity and emphasize the importance of a proper selection of the resting-state frequency range in fMRI studies on cognitive function.

## Introduction

Functional magnetic resonance imaging (fMRI) is sensitive to changes in the blood oxygen level-dependent (BOLD) signal, which originates from the neurovascular coupling, the mechanism that links neuronal activity to changes in cerebral blood flow. In particular, fMRI of the brain’s restingstate, which relies on the detection of spontaneous cerebral fluctuations, has found wide applications to study brain function in a large variety of brain conditions and disorders. Previous studies have demonstrated that the signal component of the spontaneous cerebral fluctuations, in the frequency range of 10-100 mHz, correlates with cognitive performance and thus reflects neuronal activity (1, 2). However, the dynamics of the BOLD response induced by neuronal activity is complex, not completely understood, and represents only a small part of the fMRI signal. In practice, the measured fMRI signal does not only contain the spontaneous neuronal fluctuations, but is mixed with oscillations of (non-neuronal) systemic origin and is rather noisy. Therefore, isolating the neuronal component from the BOLD signal is challenging, as there are many other non-neuronal contributions, with potentially overlapping frequency components, and even becomes more complicated in case of vascular pathology.

The primary frequencies of the cardiac (1 Hz) and respiratory (0.3 Hz) cycles contribute to the fMRI signal with higher frequencies than the frequency range pertaining to the spontaneous neuronal fluctuations, though they can appear as low-frequency fluctuations due to aliasing (3). Also systemic physiological processes such as the cerebral autoregulation, blood pressure variability, vasomotion, and the interaction between cardiac and respiratory cycles exhibit low-frequency oscillations in or close to the frequency interval of the spontaneous cerebral fluctuations (4, 5). To overcome contamination of non-neuronal signals, the raw fMRI signal is often corrected by removing correlates of cardiac and respiratory cycles (6, 7). However, to what extent the resting-state frequency component or other frequency components are associated with cardiac, respiratory, or other physiological processes remains largely unknown. In addition, changes in the vascular wall due to ageing and disease may affect the neurovascular coupling and thus also the desired neuronal signal component (8). Consequently, this complicates the interpretation of the fMRI signal and its alterations in individuals with cardiometabolic risk factors. Therefore, it is important to unravel how nonneuronal signal variations, for instance cardiac, vascular and respiratory oscillations, may contribute to, and if and how cardiometabolic risk factors have influence on the measured BOLD signal.

A commonly applied method to identify characteristic frequency components in dynamic brain signals is the Fourier-transformation (FT). However, FT assumes continuously ongoing (stationary) components. As both the neuronal and (non-neuronal) systemic physiological signals contain frequencies that vary over time (9, 10), only capturing information in the frequency domain may be insufficient. Wavelet transformation (WT) can be used to separate time-series into different frequency subbands with different time resolutions, as it can better describe non-stationary components.

The main objective of this study is to investigate to what extent the measured BOLD signal and, in particular, whether the frequency interval pertaining to the spontaneous cerebral fluctuations (10-100 mHz), is associated with various physiological influences of systemic (i.e., heart rate, autonomic function, blood pressure variability, and endothelial function) and neuronal (i.e. cognition) origin. Additionally, we investigate whether these associations were independent of cardiometabolic risk factors. The results will further improve our understanding of the interrelated neuronal and vascular contributions to the fMRI signal in the study of brain function in individuals with and without cardiometabolic risk factors.

## Methods

### The Maastricht Study: population and design

We used data from The Maastricht Study, an observational prospective population-based cohort study (11), which focuses on the etiology, pathophysiology, complications, and comorbidities of chronic diseases with extensive phenotyping. Participants were aged between 40 and 75 years, with an oversampling of individuals with type 2 diabetes. The present report includes cross-sectional data from the first 3451 participants, who completed the baseline survey between November 2010 and September 2013. MRI measurements were implemented from December 2013 onwards until February 2017 and were available in 2318 out of 3451 participants. Of the 2318 participants with available MRI measurements, 2302 subjects had complete data without artifacts, of whom 1,651 participants had complete data on physiological measurements. The study has been approved by the institutional medical ethical committee (NL31329.068.10) and the Minister of Health, Welfare and Sports of the Netherlands (Permit 131088-105234-PG). All participants gave written informed consent.

### Magnetic Resonance Imaging

Brain MRI was performed on a 3T system (Magnetom Prisma-fit Syngo MR D13D, Erlangen, Germany) by use of a 64-element head/neck coil for parallel imaging. The MRI protocol consisted of a 3D T1-weighted magnetization prepared rapid acquisition gradient echo (MPRAGE) sequence (TR/TI/TE 2300/900/2.98 ms, 176 slices, 256×240 matrix size, 1.00 mm cubic voxel size). Restingstate functional MRI (rs-fMRI) data were acquired using a task-free T_2_*-weighted blood oxygen level-dependent (BOLD) sequence (TR/TE 2000/29 ms, flip angle 90°, 32 transverse 4.00 mm thick slices, 104×104 matrix size, 2.00×2.00 mm pixel size, and 195 dynamic volumes).

Contra-indications for MRI assessments were the presence of a cardiac pacemaker or implantable cardioverter-defibrillator, neurostimulator, non-detachable insulin pump, metallic vascular clips or stents in the head, cochlear implant, metal-containing intra-uterine device, metal splinters or shrapnel, dentures with magnetic clip, an inside bracket, pregnancy, epilepsy, and claustrophobia.

### Image processing

First, affine registrations of the fMRI image to the T1 image and of the T1 image to T1 MNI-152 standard space (12) were performed. These two transformations were combined and the inverse transformation matrix was applied to the AAL2 template. T1-weighted and FLAIR images were segmented by use of a certified (ISO13485:2012), automated method (which included visual inspection) (13, 14) into white matter (WM), gray matter (GM), and cerebrospinal fluid (CSF). Details of this method are described previously (15, 16). Pre-processing of the rs-fMRI data was performed using a combination of tools in FSL 5.0.10 (FMRIB Analysis Group, University of Oxford, Oxford, U.K.) and Statistical Parametric Mapping (SPM) 12 (The Wellcome Trust College London, London, U.K.), and included magnetization stabilization followed by correction for field inhomogeneities (17), slice-timing, and head motion (18). Next, rs-fMRI data were spatially filtered to increase signal-to-noise ratio (SNR). Thereafter, the subject-specific WM and CSF masks were linearly co-registered to the rs-fMRI data using FSL’s FLIRT (19) to select only gray matter voxels for the atlas regions.

Subsequently, time-series were extracted for all gray matter voxels and these time-series were standardized by subtraction of the mean signal and dividing by the standard deviation. Finally, from each time-series, the first 128 time-points were selected. This ensures that the wavelet transformation is applied to an input signal with length 2^n^.

### Wavelet transformation

The basic idea of the stationary wavelet transform (WT) is the repeated application of high and low pass filters to a time-series, in order to obtain a multi-level representation in which each value is associated both with an interval of time and a frequency (sub)band. Each time-series was decomposed into seven wavelet subbands (Figure 1), with the Daubechies-4 wavelet chosen as the mother wavelet function (20). We applied the stationary WT, in which no subsampling is performed, hence the length of the output of each filter is equal to the length of its input. To determine to which extent the signal is structured, the relative energy per subband WE_j_ was calculated with the following formulas:

**Figure 1:**
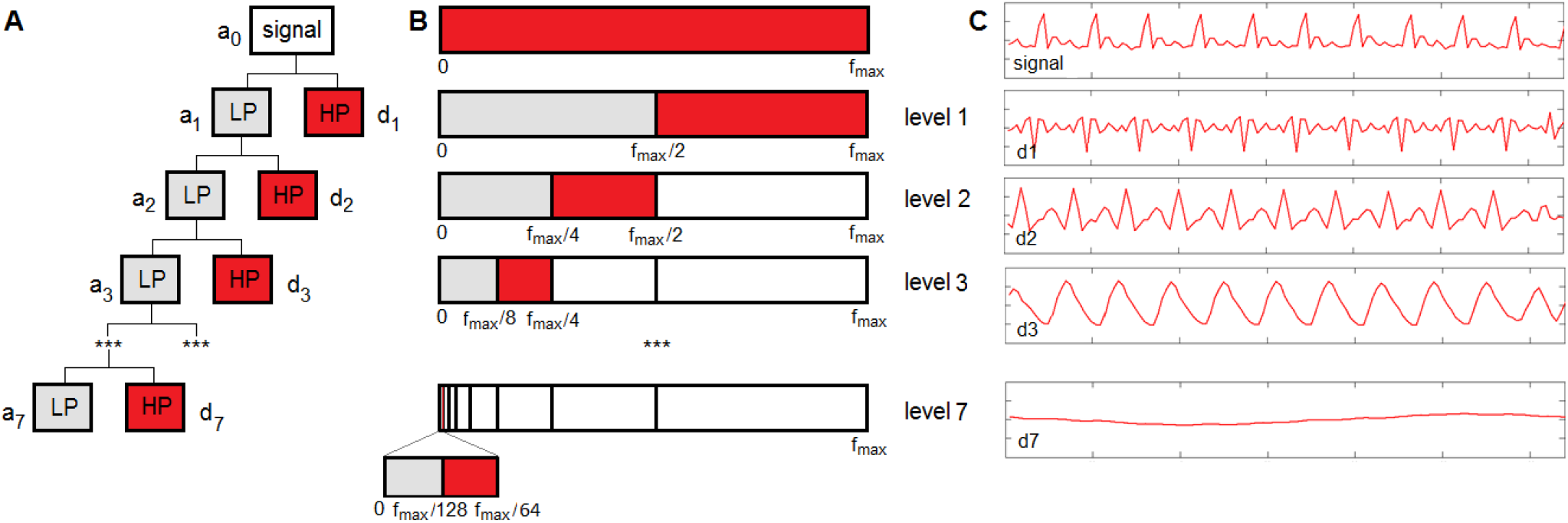
Wavelet transformation. A) Filters applied to the standardized fMRI signal. LP = low pass filter. HP = high pass filter. a_j_ and d_j_ are the scale and wavelet coefficients for subband j, respectively. B) The upper bar represents the full-standardized fMRI signal ordered by frequency. The other bars represent in gray the low frequency components and in red the high frequency components. In the remainder of the text, the HP filtered parts, i.e., the red bars, are referred to as subbands. Note that for the fMRI measurements the value of f_max_= 1/(2·TR) = 250 mHz.

The mean wavelet energy *E*_*j*_ for subband *j* is calculated by: *E*_*j*_ *=* ∑*S*_*j*_(*k*)^2^, *k*=time-point.The total energy over all the subband is: *E*_*T*_ *=* ∑*E*_*j*_. After normalization, the relative wavelet energy WE for subband *j* is: 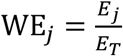

In figure 2, the frequency ranges of the wavelet subbands are schematically illustrated together with the typical frequency ranges of the physiological signals: cardiac activity (0.6-2.0 Hz), respiration (0.145-0.6 Hz), myogenic activity (0.052-0.145 Hz), neurogenic activity (0.021-0.052 Hz), and endothelial activity (0.0095-0.021 Hz) (21, 22). Note that in practice these frequency ranges overlap and are not fully adjacent or separated.

**Figure 2:**
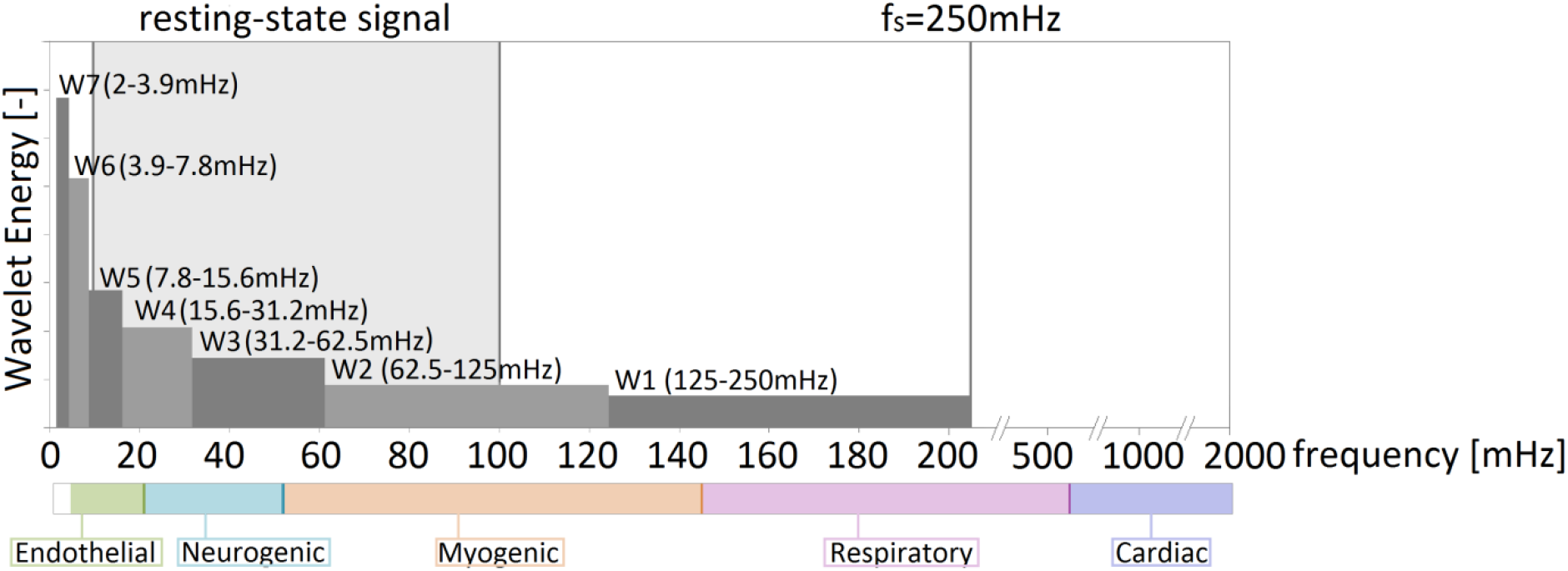
Frequency characteristics of the wavelet energy. W1-W7 indicate the frequency ranges of the wavelet subbands. On the horizontal-axis, the frequency scale is depicted. The colors indicate the subbands of the physiological signals. The gray box indicates the frequency range (10-100 mHz) for the spontaneous neuronal fluctuations; f_s_ indicates the highest frequency in the BOLD signal spectrum and is equal to half the fMRI sampling frequency.

### Physiological measures

In order to investigate the impact of physiological processes, we selected for each of the typical frequency ranges a physiological measure from the large Maastricht Study dataset. Table 1 summarizes physiological measures, their corresponding frequency ranges of the underlying physiological signal, and the overlapping wavelet subbands. The lowest physiological frequency subband, endothelial activity, captures the constant and oscillatory components of skin microvascular blood flow, which can be measured with laser-Doppler flowmetry (LDF). In the LDF data, different frequencies can be recognized, including an endothelial component. Previous studies found that the amplitude of the fluctuations in the endothelial frequency component could detect early endothelial dysfunction (23, 24). For the neurogenic subband, we used the information processing speed scores derived from cognitive tests, which is one of the main characteristic features of human cognition (25). Previous studies have already demonstrated that the resting-state frequency interval correlates with cognitive performance (1, 2). Blood pressure (BP) shows beat-to-beat oscillations due to the interplay of several cardiovascular systems, i.e., oscillations at very low (<70 mHz), low (∼100 mHz), and high (>150 mHz) frequencies, originating from myogenic vascular function, the sympathetic nervous system (Mayer waves), and endothelial-derived nitric oxide, respectively (26, 27). According to the literature, low frequency (LF) bands of blood pressure variability correlate well with measures of both sympathetic and parasympathetic activity and high frequency (HF) bands with measures of parasympathetic activity (28). As blood pressure contains information about multiple frequencies, we use blood pressure average real variability (BPV), and additionally, the LF band as a proxy for myogenic activity and the HF band for respiration (28). For the cardiac subband, we use the heart rate. At rest, the basic heart frequency is around 1 Hz, but this measure can be lower in more athletic persons, and higher (up to 1.6 Hz) in subjects with cardiovascular problems (29).

**Table 1:** Overview of the physiological measures, type of measurement, the corresponding frequency ranges and how the defined wavelet subbands overlap with the physiological frequency subbands.

| Physiological measure | Measurement | Physiological frequency subband | Wavelet subband |
| --- | --- | --- | --- |
| Endothelial component (EC) | Skin microvascular flow motion | Endothelial 9.5-21 mHz | 5 |
| Information processing speed (IPS) | Cognitive test battery | Neurogenic 21-52 mHz | 3 & 4 |
| Low frequency component Diastolic blood pressure (DiaLF) | Autonomic function test | Myogenic 52-145 mHz | 2 & 3 |
| Systolic blood pressure average real variability (BPV) | 24h ambulatory blood pressure measurement |  |  |
| High frequency component Diastolic blood pressure (DiaHF)* | Autonomic function test | Respiratory 145-600 mHz | 1 |
| Average heart rate (HR)* | 24h ambulatory blood pressure measurement | Cardiac 600-2000 mHz | 1 |

\*Note that the physiological signals with frequency components higher than half the sampling frequency (i.e. 250 mHz) become attenuated due to the low-pass signal filtering nature of the brain, but may still fold back to lower frequencies.

#### Skin microvascular flow motion

Cutaneous blood perfusion was measured by means of a laser-Doppler system (Periflux 5000; Perimed, Järfälla, Sweden), equipped with a thermostatic laser-Doppler probe (PF 457; Perimed) at the dorsal side of the wrist of the left hand as previously described (30). The LDF output was recorded for 25 min with a sample rate of 32 Hz, which gives a semi-quantitative assessment of skin microvascular blood perfusion expressed in an arbitrary perfusion unit (i.e. proportional to the product of velocity and concentration of moving red blood cells (31)). LDF skin measurements reflect perfusion in predominantly arterioles and venules (32). To quantify LDF power density Fast-Fourier transformation was performed by means of Perisoft dedicated software (PSW version 2.50; Perimed). The absolute endothelial skin microvascular flow motion component, 10– 20 mHz, was selected from the full frequency spectrum (0.01-1.6 Hz) (33).

#### Neurocognitive assessment

Cognitive performance was assessed by a concise neuropsychological test battery (11). A detailed description of neuropsychological tests and methods used to calculate domain scores is provided in the online supplementary material. Information processing speed served as the key cognitive performance measure, as its decrements are known for the current population and it involves a large part of the cerebrum. Briefly, information processing speed was derived as the average of the z-scores of the Stroop Color-Word Test Part I and II (34), the Concept Shifting Test Part A and B (35), and the Letter-Digit Substitution Test. Some of the individual test scores were log-transformed to fulfill the normality assumption and/or inverted (Stroop Color-Word Test and Concept Shifting Test) so that higher scores indicated better cognitive performance (higher information processing speed).

#### Cardiovascular autonomic function

Cardiovascular autonomic function was expressed in spectral properties of continuously measured diastolic BP. A three-lead electrocardiogram was recorded simultaneously with blood pressure using an inflatable cuff of optimal size placed around the left index finger with the Nexfin HD Monitor (BMEYE, Amsterdam, The Netherlands) (36, 37). Measurements were performed in standing position and arm position was secured using a shoulder sling to maintain blood pressure recordings at the level of the heart. In special purpose developed software, the originally recorded signals (in standing position minimally 300 beats) were converted to a format that blood pressure and electrocardiogram artifacts could be corrected for both automatically and by visual inspection. Finally, to enable spectral Fast Fourier Transform (FFT) power computations, blood pressure data files were equidistantly resampled with a 5.12 Hz frequency. Thereupon, low frequency components (<0.04 Hz) were filtered out using the smoothness prior approach (38). Finally, statistical and spectral properties of all variables were computed using Matlab routines. Spectra were computed using a 50% overlap Welch transform on data epochs of 100 seconds, allowing for a spectral resolution of 0.01 Hz. The absolute and relative power of diastolic BP in the low frequency (LF; 0.04-0.14 Hz) and the high frequency (HF; 0.15-0.4 Hz) band were quantified by spectral analysis.

#### Ambulatory 24-h blood pressure

Ambulatory 24-h blood pressure and heart rate were measured at the non-dominant arm using an ambulatory device that was programmed to take blood pressure readings every 15 minutes from 8 a.m. to 11 p.m. and every 30 minutes from 11 p.m. to 8 a.m. (Watch BP O3, Microlife, Switzerland). 24-hour BP variability was calculated as the average real variability of blood pressure readings, during 24 hours. 24-h systolic and diastolic blood pressure and heart rate were calculated as the mean of hourly averages during wake time (9 a.m. to 9 p.m.)

#### Other phenotypical measures

Educational level (low, intermediate, high), smoking status (never, current, former) and history of cardiovascular disease (CVD) were assessed by questionnaires (11). Medication use was assessed in an interview where generic name, dose, and frequency were registered. We measured weight, height, BMI, waist circumference, office blood pressure ([Omron 705IT, Japan), HbA1c, and plasma lipid profile (11). Participants were considered to have type 2 diabetes (T2DM) if they had a fasting blood glucose ≥7.0 mmol/l, or a 2-h post-load blood glucose ≥11.1 mmol/l or used oral glucose-lowering medication or insulin, and prediabetes if they had a FBG ≥6.1 mmol/l and/or a 2-h post-load blood glucose ≥7.8 mmol/l. Furthermore, participants were regarded as obese if they have BMI≥30 kg/m^2^; hypertension if their office systolic blood pressure was ≥140 mmHg and office diastolic blood pressure ≥80 mmHg or they use blood pressure lowering medication; and dyslipidemia if their totalcholesterol-to-HDL ratio was > 5 or they use lipid-modifying medication (39).

### Statistical analysis

In this explorative study, multivariable linear regression analysis was used to investigate the association of the physiological measures with the wavelet energy per subband. In these analyses, each physiological measure from Table 2 was separately included as independent variable, and the energy per subband was used as dependent variable. We discerned wavelet subbands with frequencies corresponding to the spontaneous neuronal fluctuations (10-100 mHz, i.e. subbands 2 to 5) from those beyond this range. Analyses were adjusted for age, sex, MR scanner software patch, and MRI lag time (time between physiological measures and MRI scan). Furthermore, multivariable linear regression analysis was used to investigate whether associations were independent of the four important, well-characterized cardiometabolic risk factors, highly prevalent in this population, namely T2DM (0=no, 1=yes), BMI (continuous), hypertension (0=no, 1=yes), and dyslipidemia (continuous variable total-to-HDL-cholesterol-ratio, adjusted for lipid-modifying medication) with the wavelet energy per subband. P-values < 0.05 were considered significant.

**Table 2:** General characteristics of participants according to presence of cardiometabolic risk factors.

| Characteristic | Total<br>N=1,651 | No<br>cardiometabolic<br>risk factors<br>N=535 | $\geq 1$<br>cardiometabolic<br>risk factors<br>N=1116 | P |
| --- | --- | --- | --- | --- |
| **Demographics** |  |  |  |  |
| Age (years) | 59.0±8.1 | 56.2±7.9 | 60.4±7.8 | <0.001 |
| Sex, female (%) | 48.1 | 66.5 | 39.3 | <0.001 |
| Education level (%), Low/Middle/High | 28.5/30.0/41.5 | 21.9/30.1/48.0 | 31.7/29.9/38.4 | <0.001 |
| MRI lag time (years) |  |  |  |  |
| **Cardiometabolic risk factors** |  |  |  |  |
| T2DM, yes (%) | 21.3 | 0 | 31.5 |  |
| Obesity, yes (%) | 18.3 | 0 | 27.0 |  |
| Hypertension, yes (%) | 51.3 | 0 | 75.9 |  |
| Dyslipidemia, yes (%) | 41.9 | 0 | 61.9 |  |
| **Additional cardiometabolic measures** |  |  |  |  |
| BMI (kg/m<sup>2</sup>) | 26.5±4.1 | 24.2±2.6 | 27.6±4.2 | <0.001 |
| Waist circumference (cm) | 94.2±12.6 | 86.3±9.2 | 97.9±12.3 | <0.001 |
| Office Systolic blood pressure (mmHg) | 133.8±17.0 | 121.6±10.8 | 139.6±16.3 | <0.001 |
| Office Diastolic blood pressure (mmHg) | 75.9±9.7 | 70.8±7.8 | 78.3±9.6 | <0.001 |
| 24h Systolic blood pressure, during wake time (mmHg) | 124.0±12.3 | 117.7±9.3 | 127.1±12.4 | <0.001 |
| 24h Diastolic blood pressure, during wake time (mmHg) | 79.1±10.1 | 76.9±6.5 | 80.1±7.8 | <0.001 |
| Ratio of total to HDL cholesterol (-) | 3.7±1.2 | 3.3±0.8 | 3.9±1.3 | 0.001 |
| Serum total cholesterol (mmol/L) | 5.3±1.1 | 5.6±1.0 | 5.2±1.2 | <0.001 |
| Alcohol consumption (%), none/low/high | 16.7/56.1/27.3 | 11.8/57.2/31.0 | 19.0/55.6/25.4 | <0.001 |
| Smoking status (%), never/former/current | 37.7/50.7/11.6 | 43.4/46.9/9.7 | 35.0/52.6/12.5 | 0.146 |
| HbA1c (%) | 5.78±0.77 | 5.41±0.36 | 5.95±0.85 | <0.001 |
| HbA1c (mmol/mol) | 39.6±8.4 | 35.6±3.9 | 41.6±9.2 | <0.001 |
| MMSE total score (-) | 29.1±1.1 | 29.3±1.0 | 28.9±1.2 | <0.001 |
| **Physiological measures** |  |  |  |  |
| Heart rate, during wake time (bpm) | 74.0±10.1 | 73.8±9.4 | 74.0±10.4 | 0.539 |
| Systolic BP average variability (mmHg) | 9.9±2.4 | 9.2±2.1 | 10.4±2.5 | <0.001 |
| Low frequency component diastolic BP (-) | 0.79±0.13 | 0.82±0.11 | 0.78±0.14 | <0.001 |
| High frequency component diastolic BP (-) | 0.19±0.13 | 0.16±0.11 | 0.20±0.14 | <0.001 |
| Information processing speed (SD) | 0.11 [-0.40, 0.56] | 0.33 [-0.16, 0.70] | 0.00 [-0.49, 0.47] | <0.001 |
| Endothelial component (-) | 0.83 [0.47, 1.43] | 0.83 [0.48, 1.40] | 0.83 [0.47, 1.43] | 0.218 |

Data are presented as means ± standard deviation, median and interquartile range, or percentage, and stratified for having cardiometabolic risk score 0 versus $\geq 1$ . NGM indicates normal glucose metabolism; T2DM, type 2 diabetes; HbA1c, hemoglobin A1c; BMI, body mass index; HDL, high density lipoprotein; CVA, cerebrovascular accident; MMSE, Mini-Mental State Examination; BP, blood pressure; bpm, beats per minute.

## Results

### General Characteristics of the Study Population

Table 2 shows the general characteristics of the study population according to presence of cardiometabolic risk factors. Mean age was 59±8 years (mean±SD), and 48% were women. Participants with cardiometabolic risk factors were older, more often male, and more often had a lower education level. Furthermore, they had higher BMI, waist circumference, blood pressure, and HbA1c, and were cognitively slower in the information processing speed domain.

### Association between physiological measures and wavelet energy

The energy in the wavelet subbands decreases exponentially from lower to higher frequencies, and also decreases with increasing age (p<0.05) (Fig. 3). We observed significant associations between physiological measures and the subband wavelet energies in frequency ranges for which the physiological signals and the defined wavelet subbands overlap (blue cells) and do not overlap (white cells) (Table 3).

**Table 3:**
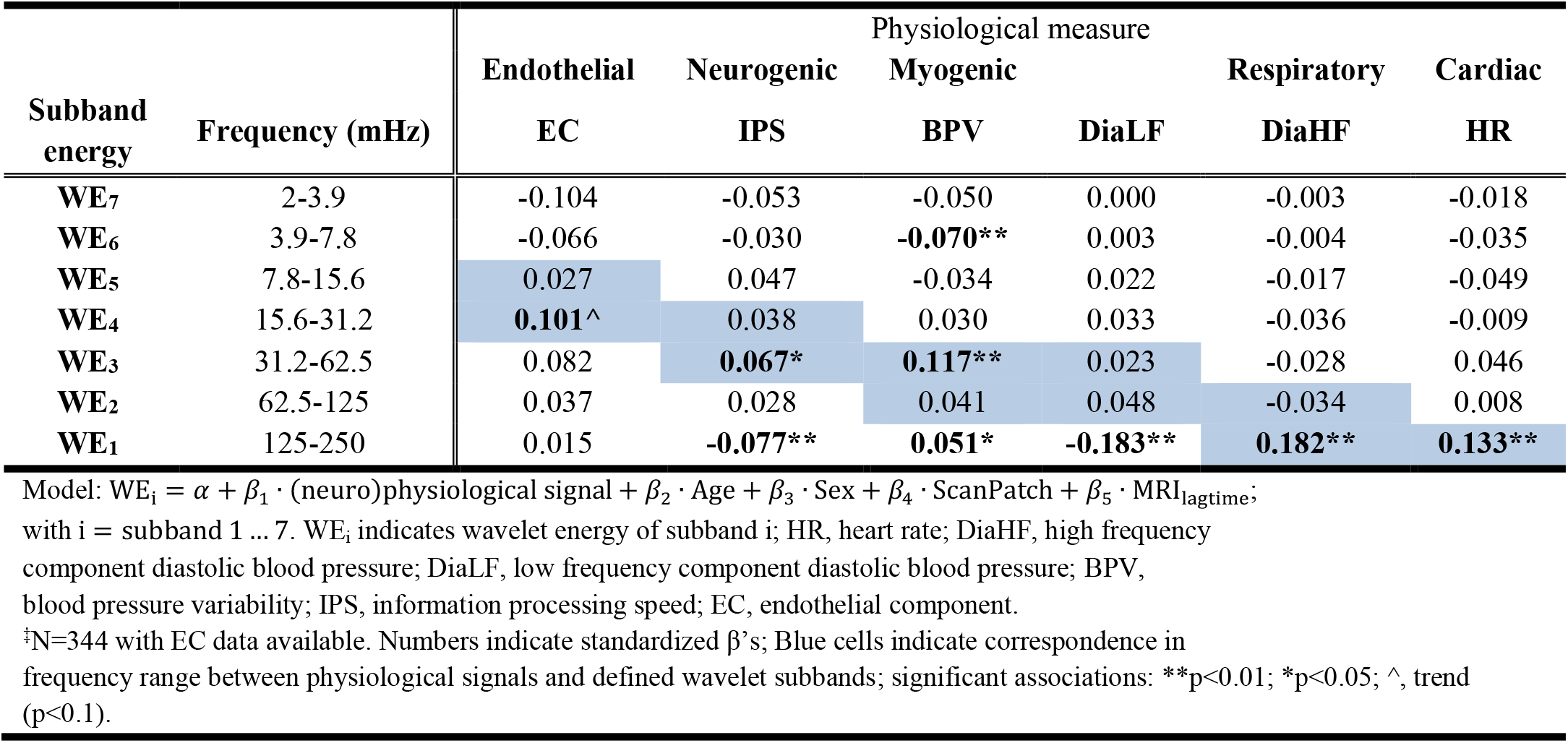
Standardized β’s for linear regression analysis of physiological measures with wavelet subband energy.

| Subband energy | Frequency (mHz) | Physiological measure |  |  |  |  |  |
| --- | --- | --- | --- | --- | --- | --- | --- |
|  |  | Endothelial | Neurogenic | Myogenic |  | Respiratory | Cardiac |
|  |  | EC | IPS | BPV | DiaLF | DiaHF | HR |
| WE<sub>7</sub> | 2-3.9 | -0.104 | -0.053 | -0.050 | 0.000 | -0.003 | -0.018 |
| WE<sub>6</sub> | 3.9-7.8 | -0.066 | -0.030 | **-0.070\*\*** | 0.003 | -0.004 | -0.035 |
| WE<sub>5</sub> | 7.8-15.6 | 0.027 | 0.047 | -0.034 | 0.022 | -0.017 | -0.049 |
| WE<sub>4</sub> | 15.6-31.2 | **0.101^** | 0.038 | 0.030 | 0.033 | -0.036 | -0.009 |
| WE<sub>3</sub> | 31.2-62.5 | 0.082 | **0.067\*** | **0.117\*\*** | 0.023 | -0.028 | 0.046 |
| WE<sub>2</sub> | 62.5-125 | 0.037 | 0.028 | 0.041 | 0.048 | -0.034 | 0.008 |
| WE<sub>1</sub> | 125-250 | 0.015 | **-0.077\*\*** | **0.051\*** | **-0.183\*\*** | **0.182\*\*** | **0.133\*\*** |

Model: $WE_i = \alpha + \beta_1 \cdot (\text{neuro})\text{physiological signal} + \beta_2 \cdot \text{Age} + \beta_3 \cdot \text{Sex} + \beta_4 \cdot \text{ScanPatch} + \beta_5 \cdot \text{MRI}_{\text{lagtime}}$ ;
with $i = \text{subband } 1 \dots 7$ . WE <sub>$i$</sub> indicates wavelet energy of subband $i$ ; HR, heart rate; DiaHF, high frequency component diastolic blood pressure; DiaLF, low frequency component diastolic blood pressure; BPV, blood pressure variability; IPS, information processing speed; EC, endothelial component.
<sup>‡</sup>N=344 with EC data available. Numbers indicate standardized $\beta$ 's; Blue cells indicate correspondence in frequency range between physiological signals and defined wavelet subbands; significant associations: \*\*p<0.01; \*p<0.05; ^, trend (p<0.1).

**Figure 3:**
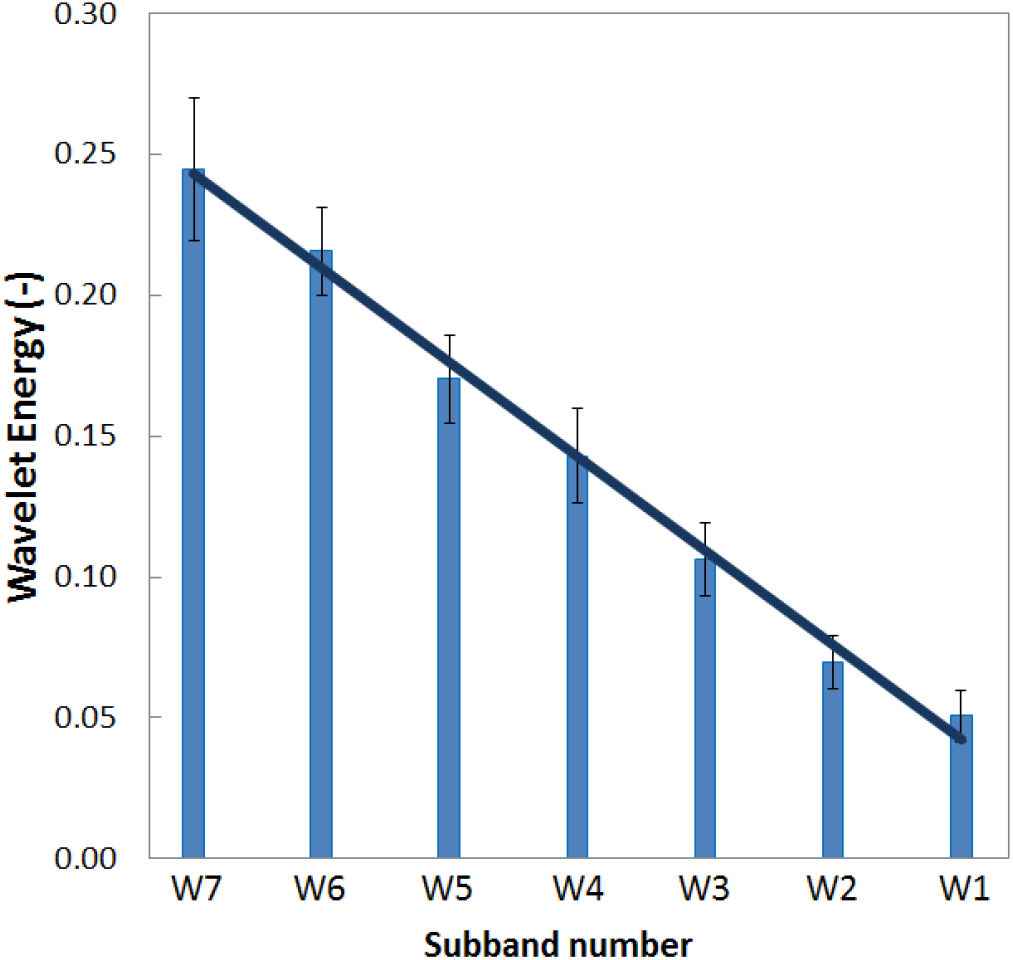
Wavelet energy of frequency subbands of the wavelet transform. Bar heights indicate mean values and error bars the standard deviations.

More specifically, in the high frequency range outside the resting-state interval (>100 mHz), all physiologic measures, except EC, were associated with the energy of subband 1 (125-250 mHz). After adjustment for the individual cardiometabolic risk factors, the association of BPV with energy of subband 1 disappeared. The association of IPS with energy of subband 1 attenuated after adjustments for hypertension and dyslipidemia, but remained significant (Supplementary Tables 1-4).

Considering the frequencies in the resting-state interval 10-100 mHz, which are commonly used for cognitive brain imaging, only the wavelet energy of subband 3 (31.262.5 mHz) was associated with IPS. (Table 1). The wavelet energies in the other subbands (2, 4 and 5) overlapping with the 10-100 mHz frequency range did not show any significant association with IPS. Also the BPV was associated with the wavelet energy for this subband 3. The observed associations were independent of having T2DM, hypertension, and dyslipidemia (Table 4 and Supplementary Table 1, 2, and 4). After adjustment for BMI the association of IPS with energy of subband 3 became stronger, and the association of BPV with energy of subband 3 weakened, but still remained significant (Table 4 and Supplementary Table 3). No significant associations with other subbands in the resting-state frequency range were found.

**Table 4:**
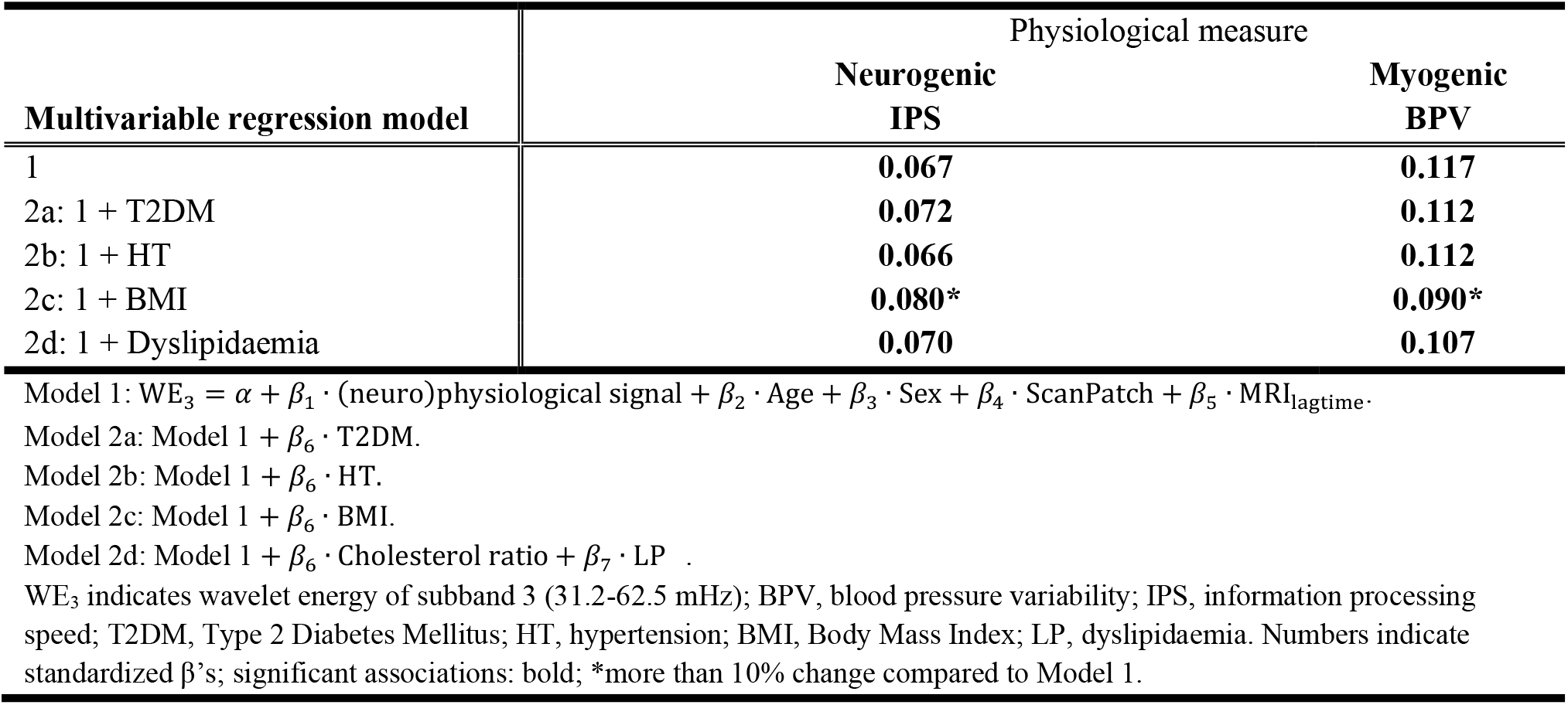
Standardized β’s for linear regression analysis of physiological measures with wavelet energy of subband 3 additionally adjusted for cardiometabolic risk factors.

| Multivariable regression model | Physiological measure |  |
| --- | --- | --- |
|  | Neurogenic<br>IPS | Myogenic<br>BPV |
| 1 | **0.067** | **0.117** |
| 2a: 1 + T2DM | **0.072** | **0.112** |
| 2b: 1 + HT | **0.066** | **0.112** |
| 2c: 1 + BMI | **0.080\*** | **0.090\*** |
| 2d: 1 + Dyslipidaemia | **0.070** | **0.107** |

Model 1: $WE_3 = \alpha + \beta_1 \cdot (\text{neuro})\text{physiological signal} + \beta_2 \cdot \text{Age} + \beta_3 \cdot \text{Sex} + \beta_4 \cdot \text{ScanPatch} + \beta_5 \cdot \text{MRI}_{\text{lagtime}}$ .
Model 2a: Model 1 + $\beta_6 \cdot \text{T2DM}$ .
Model 2b: Model 1 + $\beta_6 \cdot \text{HT}$ .
Model 2c: Model 1 + $\beta_6 \cdot \text{BMI}$ .
Model 2d: Model 1 + $\beta_6 \cdot \text{Cholesterol ratio} + \beta_7 \cdot \text{LP}$ .
WE<sub>3</sub> indicates wavelet energy of subband 3 (31.2-62.5 mHz); BPV, blood pressure variability; IPS, information processing speed; T2DM, Type 2 Diabetes Mellitus; HT, hypertension; BMI, Body Mass Index; LP, dyslipidaemia. Numbers indicate standardized $\beta$ 's; significant associations: bold; \*more than 10% change compared to Model 1.

In the frequencies lower than the resting-state interval (<10 mHz), the BPV was associated with the energy of subband 6 (3.9-7.8 mHz). Additional adjustment for cardiometabolic risk factors did not materially change the associations of the energy of subband 2, and 4-7.

## Discussion

### Current findings

We set out to find different physiological signal components in the measured resting-state fMRI signal in a large population with variations in cardiometabolic risk factors. In this study, we observed that physiological measures are associated with certain frequency subbands of the dynamic BOLD signal. Most importantly, cognitive performance was associated with the energy of frequency subband 3 (31.2-62.5 mHz), reflecting the level of neurogenic activity. However, blood pressure variability, reflecting systemic myogenic activity, was also associated with the energy of the same frequency subband. Also other physiological measures were associated with specific frequency subbands, i.e., cardiac and respiratory activity with high frequencies (>125 mHz), and endothelial and metabolic activity with low frequencies (<10 mHz).

Furthermore, we investigated whether these associations were independent of cardiometabolic risk factors. We found some associations were dependent on cardiometabolic risk factors and caution should be made when studying the resting-state fMRI signal in populations with cardiometabolic risk factors.

### Neurogenic associations

We found the only association of cognitive performance, i.e. neurogenic activity, with the energy of subband 3 (31.2-62.5 mHz). A previous study on wavelet correlation analysis observed significant associations between wavelet coefficients of the subband representing the (overlapping) 10–120 mHz frequency range and structural connectivity measures (global efficiency) (41). Another study, in which a Hilbert-Huang Transform was used, found that the corresponding frequency band 45-87 mHz was associated with cognitive function in a large aging cohort (42). Previous studies also showed that low frequency oscillations of the arterial pressure, also called Mayer waves, are positively correlated with low frequency oscillations (∼100 mHz) in the resting-state signal (4, 43).

We also found an association between the oscillations related to the (average real systolic) blood pressure variability and the energy of the neurogenic subband (wavelet subband 3, 31.2-62.5 mHz). This observation may be due to the influence of systemic blood pressure variations on the brain’s vascular system, but in part may also reflect the myogenic aspect of the neurovascular coupling, i.e. the activity of smooth muscle cells for the vasoconstriction and vasodilation of the brain vascular system to alter the local blood flow in response to neuronal activity. These cardiometabolic risk factors affect the cerebral vasculature, leading to abnormal patterns of vasodilation and vasoconstriction to facilitate the required blood flow, and also impairs the neurovascular coupling (8, 44, 45, 46). Similarly, the contribution of myogenic response is often altered due to impairment of vascular reactivity and arterial stiffness in participants with cardiometabolic risk factors (47, 48). The association of IPS with energy of subband 3 became stronger, and the association of BPV with energy of subband 3 weakened, but both still remained significant, after adjustment for BMI.

### High frequency associations

As cardiac cycles have a relatively high frequency (600-2000 mHz) compared to the low frequency (< 100 mHz) BOLD fluctuations, one would initially expect that these could be distinguished easily from the resting-state signal. However, due to the long repetition time (i.e., low sampling frequency, 250 mHz), cardiac modulations at the primary frequency (600-2000 mHz) are aliased into low frequency subbands (5, 49). A recent study observed that physiological low frequency oscillations (systemic signals without clear origin that travel with the blood), instead of aliased cardiac and respiratory signals, are most influential in the resting-state signal (50). In the present study, we also found strong associations of cardiac and respiratory signals with energy in the high frequency subband 125-250 mHz. However, we did not obtain strong associations of these signals in the lower frequency subbands, indicating that these were possibly less intertwined with the resting-state signal as expected, or highly attenuated due to the low-pass filter nature of the hemodynamic process. The association of BPV with energy of subband 1 disappeared after adjustment for all individual cardiometabolic risk factors. The association of IPS with energy of subband 1 attenuated, but remained significant after adjustment for hypertension and dyslipidemia, but disappeared when adjusted for T2DM or BMI (51).

### Low frequency associations

For frequencies lower than those corresponding to the resting-state interval (10-100 mHz), blood pressure variability (BPV) was associated with the energy of the low frequency subband 3.9-7.8 mHz, which likely reflects endothelial activity, originating from endothelial-derived nitric oxide (26, 27). T2DM and obesity were associated with the energy of the low frequency subband 5 and 6 (3.9-15.6 mHz), and dyslipidemia only with the energy of subband 5 (7.8-15.6 mHz). Previous studies also found differences in the amplitude of the BOLD signal in the time course of the hemodynamic response function, and disturbed neurovascular coupling in apparently normal brains of subjects with T2DM (52, 53). This is not surprising, because the regional cerebral blood flow, on which the BOLD response depends, increases in proportion to glucose consumption (54). Similar changes were also found in studies on participants with cerebrovascular disease (55, 56).

### Implications for clinical studies

A purer neurogenic signal for resting-state fMRI analysis, less influenced by non-neuronal physiological signals, could be obtained using only the frequency band 31.2-62.5 mHz. In addition, for the selection of the resting-state range one should also keep in mind the characteristics of the study population. The association of physiologic measures with the measured BOLD signal was found to be partly dependent on cardiometabolic risk factors. Given the fact that systemic variation in blood pressure and dilatation/constriction of blood vessels are altered in individuals with cardiometabolic risk factors, it is important to be cautious in the interpretation of changes in the measured neurogenic BOLD signal. These changes may either be due to systemic circulatory or cerebrovascular effects, the coupling between the vascular and neuronal activity or a combination of systemic and cerebral effects. The current study suggests that in order to obtain a more native signal of the spontaneous neuronal fluctuations (extracranial) blood pressure variations could be taken into account. Lastly, the direct influences of cardiac and respiratory motion are mainly recognizable in the higher frequencies (>100 mHz), and therefore possibly less influential for the resting-state interval.

### Strengths and limitations

Strengths of this study are the large sample size, the population-based design, and the extensive phenotyping, which gave us the oppurtunity to investigate a variety of systemic physiological signals relevant for the selected frequency subbands. In this way, we could show that the association between cognitive performance and the wavelet energy of subband 3 (31.2-62.5 mHz) was independent of cardiometabolic risk factors. Furthermore, the large number of rs-fMRI scans were semi-automatically processed blinded to participant characteristics, which ensures an objective analysis. There are also some limitations. First, the rs-fMRI scan protocol used in this study was designed for the large population-based Maastricht Study, and not for this very specific type of analysis. Therefore, a commonly used repetition time of 2,000 ms was chosen. However, ideally, to filter out cardiac and respiratory signals, scans with a shorter repetition time (and thus higher sampling frequency) would have been optimal. Furthermore, we did not acquire the physiological signals simultaneously with the fMRI acquisition. Finally, the reported associations between the fMRI signal components, physiological measures, and the cardiometabolic risk factors should not be interpreted in terms of formal causal relations of biological processes.

## Conclusion

We found that both neuronal and systemic non-neuronal physiological measures are associated with frequency specific subbands of the measured dynamic fMRI signal. Most importantly, the subband analysis showed an association of cognitive performance as well as blood pressure variations with the signal of the subband 3 with the frequency range 31.2-62.5 mHz, which is usually thought to reflect the signal of spontaneous neuronal fluctuations. These observations can be explained on the one hand by the intrinsic coupling between neuronal activity and the required vasoconstrictive/vasodilatory actions for the alterations in blood flow, and on the other hand by the influence of systemic blood pressure variations that also occur in the cerebral circulation. Though strong associations of primary cardiac and respiratory measures with the energy of high frequency subbands (>125 mHz) were obtained, they did not influence the energy of subbands in the interval (10-100 mHz) relevant for neuronal processes. Finally, not all associations between physiological signals and subband energies were independent of cardiometabolic risk factors, consistent with what was expected. Our findings highlight the strong intertwining of neuronal and cardiometabolic activity, and emphasize the importance of a proper selection of the resting-state frequency range. Therefore, cautiousness is advised for the interpretation fMRI signal changes as pure neuronal signals, especially in the presence of cardiometabolic risk factors.

## Data Availability

All data produced in the present study are available upon a reasonable request to management of the Maastricht Study

