## Supplementary material for "Exploring the neuronal and systemic physiological contributions to spontaneous cerebral fluctuations – Insights from functional MRI in The Maastricht Study"

**Supplementary Table 1:** Standardized β's for linear regression analysis of physiological measures with wavelet subband energy additionally adjusted for having type 2 diabetes.

| Subband energy | Frequency (mHz) | Physiological measure<br>Endothelial<br>EC | <br>Neurogenic<br>IPS | <br>Myogenic<br>BPV | <br><br>DiaLF | <br>Respiratory<br>DiaHF | <br>Cardiac<br>HR |
|---|---|---|---|---|---|---|---|
| **$WE_7$** | 2-3.9 | -0.099 | -0.054 | -0.049 | -0.004 | 0.001 | -0.015 |
| **$WE_6$** | 3.9-7.8 | -0.067 | -0.040 | **-0.065*** | -0.011 | 0.010 | -0.024 |
| **$WE_5$** | 7.8-15.6 | 0.022 | 0.034 | -0.027 | 0.006 | -0.002 | -0.037 |
| **$WE_4$** | 15.6-31.2 | **0.104^** | 0.038 | 0.032 | 0.033 | -0.035 | -0.010 |
| **$WE_3$** | 31.2-62.5 | 0.076 | **0.072**** | **0.112**** | 0.034 | -0.037 | 0.038 |
| **$WE_2$** | 62.5-125 | 0.030 | 0.028 | 0.040 | 0.047 | -0.033 | 0.007 |
| **$WE_1$** | 125-250 | 0.027 | -0.045 | 0.023 | **-0.138**** | **0.137**** | **0.105**** |

Model: $WE_i = \alpha + \beta_1 \cdot \text{(neuro)physiological signal} + \beta_2 \cdot \text{Age} + \beta_3 \cdot \text{Sex} + \beta_4 \cdot \text{ScanPatch} + \beta_5 \cdot \text{MRI}_{\text{lagtime}} + \beta_6 \cdot T2DM$; with i = subband 1 ... 7. $WE_i$ indicates wavelet energy of subband i; HR, heart rate; DiaHF, high frequency component diastolic blood pressure; DiaLF, low frequency component diastolic blood pressure; SysBP, systolic blood pressure; IPS, information processing speed; EC, endothelial component; T2DM, type 2 diabetes mellitus.
‡N=344 with EC data available. Numbers indicate standardized β's; Blue cells indicate correspondence in frequency range; significant associations: **p<0.01; *p<0.05; ^, trend (p<0.1).

**Supplementary Table 2:** Standardized β's for linear regression analysis of physiological measures with wavelet subband energy additionally adjusted for having hypertension.

| Subband energy | Frequency (mHz) | Physiological measure<br>Endothelial<br>EC | <br>Neurogenic<br>IPS | <br>Myogenic<br>BPV | <br><br>DiaLF | <br>Respiratory<br>DiaHF | <br>Cardiac<br>HR |
|---|---|---|---|---|---|---|---|
| **$WE_7$** | 2-3.9 | -0.099 | -0.051 | **-0.054*** | -0.001 | -0.002 | -0.017 |
| **$WE_6$** | 3.9-7.8 | -0.063 | -0.031 | **-0.068*** | -0.001 | 0.000 | -0.034 |
| **$WE_5$** | 7.8-15.6 | 0.027 | 0.041 | -0.024 | 0.016 | -0.012 | -0.048 |
| **$WE_4$** | 15.6-31.2 | **0.102^** | 0.035 | 0.040 | 0.030 | -0.032 | -0.009 |
| **$WE_3$** | 31.2-62.5 | 0.076 | **0.066*** | **0.112**** | 0.027 | -0.030 | 0.044 |
| **$WE_2$** | 62.5-125 | 0.033 | 0.026 | 0.043 | 0.043 | -0.030 | 0.008 |
| **$WE_1$** | 125-250 | 0.014 | **-0.065*** | 0.027 | **-0.160**** | **0.159**** | **0.132**** |

Model: $WE_i = \alpha + \beta_1 \cdot \text{(neuro)physiological signal} + \beta_2 \cdot \text{Age} + \beta_3 \cdot \text{Sex} + \beta_4 \cdot \text{ScanPatch} + \beta_5 \cdot \text{MRI}_{\text{lagtime}} + \beta_6 \cdot HT$; with i = subband 1 ... 7. $WE_i$ indicates wavelet energy of subband i; HR, heart rate; DiaHF, high frequency component diastolic blood pressure; DiaLF, low frequency component diastolic blood pressure; SysBP, systolic blood pressure; IPS, information processing speed; EC, endothelial component; HT, hypertension.
‡N=344 with EC data available. Numbers indicate standardized β's; Blue cells indicate correspondence in frequency range; significant associations: **p<0.01; *p<0.05; ^, trend (p<0.1).

**Supplementary Table 3:** Standardized β's for linear regression analysis of physiological measures with wavelet subband energy additionally adjusted for BMI.

| Subband Energy | Frequency (mHz) | Physiological measure<br>Endothelial<br>EC | <br>Neurogenic<br>IPS | <br>Myogenic<br>BPV | <br><br>DiaLF | <br>Respiratory<br>DiaHF | <br>Cardiac<br>HR |
|---|---|---|---|---|---|---|---|
| **$WE_7$** | 2-3.9 | **-0.107^** | **-0.056*** | -0.043 | -0.006 | 0.003 | -0.012 |
| **$WE_6$** | 3.9-7.8 | -0.076 | -0.040 | **-0.051^** | -0.010 | 0.008 | -0.023 |
| **$WE_5$** | 7.8-15.6 | 0.017 | 0.022 | 0.007 | -0.004 | 0.005 | -0.023 |
| **$WE_4$** | 15.6-31.2 | **0.103^** | 0.033 | 0.040 | 0.028 | -0.030 | -0.004 |
| **$WE_3$** | 31.2-62.5 | 0.086 | **0.080**** | **0.090**** | 0.040 | -0.042 | 0.027 |
| **$WE_2$** | 62.5-125 | 0.041 | 0.038 | 0.022 | **0.057*** | -0.042 | -0.005 |
| **$WE_1$** | 125-250 | 0.049 | -0.030 | 0.031 | **-0.128**** | **0.131**** | **0.087**** |

Model: $\mathrm{WE_i} = \alpha + \beta_1 \cdot \mathrm{(neuro)physiological\ signal} + \beta_2 \cdot \mathrm{Age} + \beta_3 \cdot \mathrm{Sex} + \beta_4 \cdot \mathrm{ScanPatch} + \beta_5 \cdot \mathrm{MRI_{lagtime}} + \beta_6 \cdot BMI$; with i = subband 1 ... 7. $WE_i$ indicates wavelet energy of subband i; HR, heart rate; DiaHF, high frequency component diastolic blood pressure; DiaLF, low frequency component diastolic blood pressure; SysBP, systolic blood pressure; IPS, information processing speed; EC, endothelial component; BMI, body mass index. ‡N=344 with EC data available. Numbers indicate standardized β's; Blue cells indicate correspondence in frequency range; significant associations: **p<0.01; *p<0.05; ^, trend (p<0.1).

**Supplementary Table 4:** Standardized β's for linear regression analysis of physiological measures with wavelet subband energy additionally adjusted for total-to-HDL-cholesterol-ratio and lipid-modifying medication.

| Subband Energy | Frequency (mHz) | Physiological measure<br>Endothelial<br>EC | <br>Neurogenic<br>IPS | <br>Myogenic<br>BPV | <br><br>DiaLF | <br>Respiratory<br>DiaHF | <br>Cardiac<br>HR |
|---|---|---|---|---|---|---|---|
| **$WE_7$** | 2-3.9 | -0.096 | -0.049 | -0.050 | 0.005 | -0.007 | -0.018 |
| **$WE_6$** | 3.9-7.8 | -0.060 | -0.034 | **-0.065*** | 0.001 | -0.002 | -0.029 |
| **$WE_5$** | 7.8-15.6 | 0.028 | 0.032 | -0.021 | 0.012 | -0.009 | -0.033 |
| **$WE_4$** | 15.6-31.2 | **0.101^** | 0.035 | 0.030 | 0.025 | -0.028 | -0.009 |
| **$WE_3$** | 31.2-62.5 | 0.070 | **0.070**** | **0.107**** | 0.022 | -0.026 | 0.035 |
| **$WE_2$** | 62.5-125 | 0.027 | 0.025 | 0.041 | 0.040 | -0.027 | 0.009 |
| **$WE_1$** | 125-250 | 0.015 | **-0.053*** | 0.033 | **-0.156**** | **0.155**** | **0.114**** |

Model: $\mathrm{WE_i} = \alpha + \beta_1 \cdot \mathrm{(neuro)physiological\ signal} + \beta_2 \cdot \mathrm{Age} + \beta_3 \cdot \mathrm{Sex} + \beta_4 \cdot \mathrm{ScanPatch} + \beta_5 \cdot \mathrm{MRI_{lagtime}} + \beta_6 \cdot Cholesterolratio + \beta_7 \cdot LP_{med}$; with i = subband 1 ... 7. $WE_i$ indicates wavelet energy of subband i; HR, heart rate; DiaHF, high frequency component diastolic blood pressure; DiaLF, low frequency component diastolic blood pressure; SysBP, systolic blood pressure; IPS, information processing speed; EC, endothelial component; $LP_{med}$, lipid-modifying medication. ‡N=344 with EC data available. Numbers indicate standardized β's; Blue cells indicate correspondence in frequency range; significant associations: **p<0.01; *p<0.05; ^, trend (p<0.1).
